# Simulation of synthetic health records for assessment of causal inference methods for vaccine efficacy

**DOI:** 10.64898/2026.07.17.26358308

**Authors:** Víctor Velasco-Pardo, Luke Daines, S. Vittal Katikireddi, Lewis Ritchie, Chris Robertson, Colin R. Simpson, Colin McCowan, Ben Swallow

## Abstract

**Background:** During the COVID-19 pandemic, public health agencies used near real-time observational data to answer questions regarding vaccine effectiveness. However, traditional observational methods do not allow conclusions regarding counterfactual scenarios to be drawn from clinical data. Counterfactuals, which are outcomes that would have occurred under alternative interventions, can be used to formally assess the causal effects of public health interventions on health outcomes while accounting for the effects of confounding. Ideally individual patient data is used for the development of counterfactuals. Low-fidelity synthetic data may be useful for advancing methodological development where governance and privacy constraints prohibit access to sensitive personal data.

**Methods:** We simulated synthetic datasets based on the EAVE-II COVID-19 platform which has been limited to use for surveillance purposes. EAVE-II includes almost all resident people in Scotland registered with qualified general medical practitioners. Patient characteristics were simulated to reflect the known distribution of the Scottish population, accounting for dependencies between variables. Each synthetic dataset was encoded to different realistic scenarios for EAVEII ‘ground truth’ vaccine rollout and effectiveness results, explicitly stating the causal and confounding mechanisms, using a statistically sound method based on marginal structural models. Synthetic datasets of 100,000 individuals were then generated across five confounding scenarios and five severe outcome types.

**Results:** In scenarios with weak confounding, both unweighted and inverse probability of treatment weighted (IPTW) logistic regression recovered the true causal parameters. As confounding strength increased, only weighted models recovered the true mechanism.

**Conclusions:** Low-fidelity synthetic datasets simulated from EAVE-II data analysts to build and test causal inference pipelines, develop novel analysis pipelines, and train new researchers while awaiting access to real data. We showed how to generate synthetic datasets from a marginal structural model under different confounding scenarios.

## Introduction

In Phase 3 clinical trials, efficacy of SARS-CoV-2 vaccines was demonstrated against severe disease, hospitalisation, and death resulting from COVID-19-related complications (such as acute respiratory distress syndrome, thrombotic events, or secondary organ failure (1–3)). Observational data collected from electronic health records (EHRs) in a real-world setting can be complimentary to post-Phase 3 clinical trials. Observational studies can investigate real-world vaccine effectiveness and safety (4). Important questions to inform the roll out of vaccination programmes relate to the relative effectiveness of different vaccine schedules, the efficacy of heterologous vaccine schedules (5,6), and the potential effect of SARS-CoV-2 vaccines on severe clinical outcomes from other respiratory infections, such as increased susceptibility to influenza, pneumonia, or exacerbations of chronic respiratory disease (7).

Ideally, causal questions determining the correct pathway and treatment effects regarding vaccines and other medical interventions are answered by means of randomised controlled trials (RCTs). When it is not possible to perform an RCT, due to time constraints, economic or ethical reasons, EHRs can be used to emulate a clinical trial (8,9). Furthermore, in a pandemic setting observational studies using data from electronic health records can be undertaken to address any knowledge gaps not addressed through clinical trials. For example, phase 3 clinical trials were underpowered for AstraZeneca Oxford mRNA vaccine in older people (10). To infer causality and remove the effect of confounders, such as age, ethnicity, and comorbidities, we would ideally use formal causal inference methods, explicitly modelling potential outcomes (11). Developing new methodological tools is challenging due to privacy and confidentiality constraints: researchers commonly only gain access to databases with individual patient data with agreed permissions, for necessary analysis rather than specific methodology development.

In general, it is common to use synthetic data to assess the performance of existing and newly developed statistical methods. In the context of public health databases such as EAVE-II, synthetic data also allow developers to develop new methods and pipelines before access to the data has been granted, and importantly without compromising privacy (12). Having a simulated version of the requested data available means that initial code can be developed even while waiting for data access to be granted.

The EAVE-II data platform reporting on the Scottish population, the protocol of which is outlined by Simpson et al. (13), was developed to facilitate the growing need for rapid, large observational epidemiological studies to identify the epidemiological and population-level clinical effects of the COVID-19 pandemic. The platform links different health databases and contains patient characteristics, as well as events such as vaccine uptake, serology data, diagnostic tests (RT-PCR), hospitalisations and deaths, allowing the EAVE-II team to conduct observational studies in response to public health questions. The linked dataset contains records for 5.4 million people, almost the entire Scottish population (14). Using this platform, the EAVE-II group generated evidence to answer key public health questions during the COVID-19 pandemic (7,15–17), as well as comparative studies of vaccination schedules in international contexts (6). However, these papers could only report associations as causal inference methods were not implemented.

In this study, we used the EAVE-II platform to assess the effectiveness of different COVID-19 vaccine schedules in preventing hospitalisation and death and whether the primary schedule of COVID-19 vaccines had an effect on the effectiveness of winter influenza vaccinations in the Scottish population. To allow us to accelerate the research process, remove the effect of confounders, and undertake a causal inference study, but lacking access to individual patient data, we aimed to create realistic scenarios using synthetic data and then perform our analysis. The demographic characteristics of the synthetic individuals, which will inform both treatment allocation and outcomes, should reflect their distribution in the Scottish population.

## Methods

### Causal inference background

The fundamental problem of causal inference as originally described by Holland (18), is the impossibility of observing all potential outcomes in the same patient. If Y(0) and Y(1) denote the clinical outcome after treatment A = 0 or A = 1 is given, respectively, only one of the two potential outcomes can be observed in each patient, as they cannot be allocated both treatment and control simultaneously. The goal of causal inference methods is to infer the effect of a treatment on an outcome adjusting for confounders. The validity of conclusions reached through causal inference methods relies on assumptions (11,19) including positivity (any treatment group has a positive probability of receiving any treatment), consistency (the potential outcome for the observed treatment equals the observed outcome), and exchangeability (all confounders have been measured).

We introduce the setting and notation used throughout this paper, following Seaman and Keogh (30). We consider K + 1 visits to the clinic, indexed by k = 0, …, K. For each patient, A = (A_0_, …, A_K−1_) denotes treatment allocations, Y = (Y_1_, …, Y_K_) represents observed outcomes, and Z = (Z_1_, …, Z_q_) denotes the confounding variables that influence both treatment allocation and outcomes. Each Yₖ indicates the outcome observed on the kth visit, and each Aₖ indicates the treatment allocated on that visit after Yₖ was observed. In the applied setting considered here, the variables recorded in Z are sex and age.

### Marginal structural models

We focus attention on marginal structural models (MSMs (20,21)). These are generalised regression models of the outcome of interest Y on the treatment A. Marginal structural models provide a flexible approach to causal modelling, allowing for both time-varying treatments and time-dependent confounders. They specify a parametric model of the marginal effect of treatment A on outcome Y:

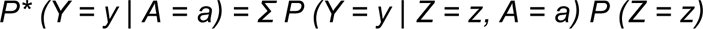

where Z denotes the vector of confounders. The effect of the confounders is marginalised out, and the resulting model describes the relationship between the outcome Y and the treatment A. A link function g() may be set to the identity, logarithm, logistic, or cloglog function for linear, count, binary, and survival responses respectively. The assumptions regarding the direction of causality are illustrated in Figure 1 using a directed acyclic graph (DAG). The confounders Z influence every other variable in the DAG, and treatments and outcomes influence one another in the sequential manner indicated.

**Figure 1.**
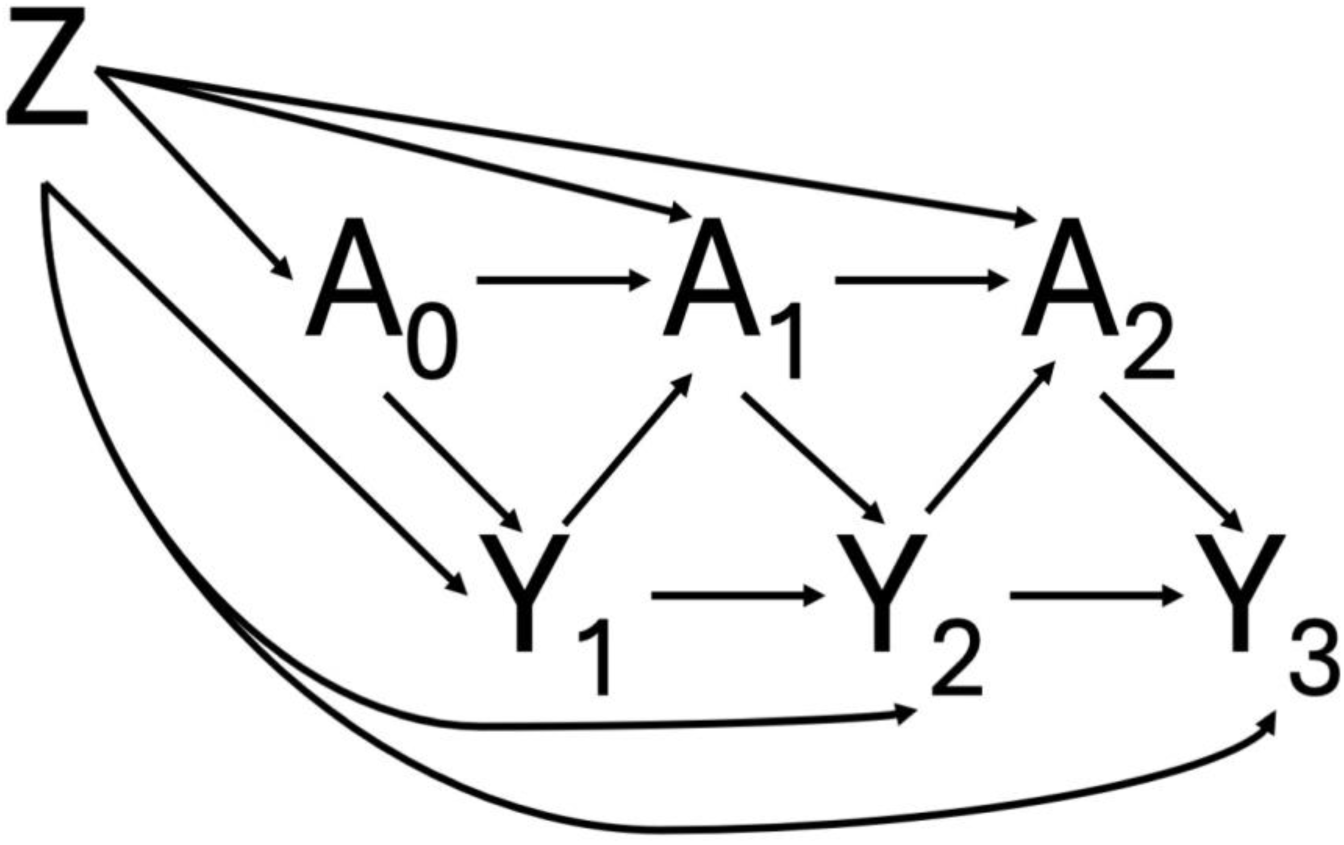
Directed acyclic graph (DAG) illustrating the causal assumptions for a marginal structural model with four clinic visits (k = 0, 1, 2, 3). Z = confounders; Aₖ = treatment allocation at visit k; Yₖ = outcome at visit k.

In an RCT setting, randomisation ensures that the causal effect is correctly estimated as the confounders are balanced out. In an observational study it is necessary to adjust for confounding using appropriate statistical methods. The adjustment for confounding typically involves ’weighting’ observations to account for the joint distribution of treatment allocation and confounders, such that observations with a treatment regime uncommon for their observed confounding values are ’weighted up’ and those with a more typical treatment regime are ’weighted down’.

Alternatively, ’matching’ observations across treatment groups who have similar confounding characteristics can be used to create this balanced joint distribution.

### Parameter estimation and propensity scores

Causal inference methods weight or match observations according to their ’propensity’ to be allocated a certain treatment given their values for the confounding variables. That propensity is quantified by the ’propensity score’ (11) which for a binary treatment variable A and a vector of q confounders Z = (Z_1_, …, Z_q_) is defined as e(Z) = E(A|Z) = P (A = 1|Z). For high-dimensional Z, a binary propensity score is assumed to follow a logistic model of the confounders and can therefore be estimated via logistic regression. Li and Li (22) generalised the concept of propensity score to multiple treatments: if a treatment variable A can take values j = 1, …, J, the generalised propensity score can be defined as eⱼ(Z) = P (A = j | Z). The most common method for fitting MSMs is inverse probability of treatment weighting (IPTW), where the MSM is fitted using estimated propensity scores to weight observations (20,23).

### Motivation for synthetic data

(12)We use a statistically sound approach to simulate data where the causal and confounding mechanisms are stated explicitly, allowing us to test the performance of causal inference methods against the synthetic data. The problem of simulating synthetic data from ’ground truth’ causal models (where the effect sizes have known values) has been addressed by a number of authors (24–30). In what follows, we focus on the approach of Seaman and Keogh (30) in simulating causally motivated data.

### Data simulation framework

Seaman and Keogh (30) substantially extended the framework of Evans and Didelez (29). In that approach, the joint probability distribution is specified by means of the following four components: (1) the distribution of confounders, P(Z = z); (2) the causal effect of treatment on the outcome, P*(Y = y | A = a); (3) a scalar ’risk score’ function h(Z, Āₖ) that ranks individuals according to confounder-driven risk; and (4) an association parameter −1 < ρₖ ≤ 0, determining the strength of dependence between h(Z) and Y. When ρₖ = 0 the confounders do not influence the outcome Y, and when ρₖ ≈ −1 the confounding mechanism is strongest.

The only drawback of this approach compared to that of Evans and Didelez (29) is the lack of a readily available package implementing the simulation algorithm. Therefore, the data simulation algorithm needs to be implemented by the practitioner. We implemented the algorithm outlined by Seaman and Keogh (30), §3.3 and Appendix D, in R. Code to produce the results in this paper is available at https://github.com/victorvelasco/eave_ii_simulated_data. The pseudocode is reproduced in Appendix A.

### Confounders

We consider sex and age as confounders, encoded as a binary vector Z = (Z_1_, …, Z_16_). Sex is coded as Z_1_ = 1(Sex = M). Age is an ordinal variable with sixteen categories, encoded following Harrell (31) §2.7.3) as Z₂ = 1(Age ≥ 15), …, Z_16_ = 1(Age ≥ 90). To ensure the simulated Z reflects population-level patterns in Scotland, we used aggregated counts from EAVE-II extracted in August 2025, stratified by sex and age group. Baseline characteristics of the EAVE-II population (N = 4,567,266) are shown in Table 1.

**Table 1.** Baseline characteristics of the EAVE-II population that is used to simulate synthetic datasets (N = 4,567,266).

|  | Female (N=2312118) | Male (N=2255148) | Total (N=4567266) |
| --- | --- | --- | --- |
| **Age group (years)** |  |  |  |
| 15-20 | 130283 (5.6%) | 138830 (6.2%) | 269113 (5.9%) |
| 20-25 | 153898 (6.7%) | 150748 (6.7%) | 304646 (6.7%) |
| 25-30 | 182148 (7.9%) | 178247 (7.9%) | 360395 (7.9%) |
| 30-35 | 189079 (8.2%) | 196734 (8.7%) | 385813 (8.4%) |
| 35-40 | 183416 (7.9%) | 193998 (8.6%) | 377414 (8.3%) |
| 40-45 | 172465 (7.5%) | 184932 (8.2%) | 357397 (7.8%) |
| 45-50 | 165628 (7.2%) | 174722 (7.7%) | 340350 (7.5%) |
| 50-55 | 194211 (8.4%) | 195635 (8.7%) | 389846 (8.5%) |
| 55-60 | 200483 (8.7%) | 197031 (8.7%) | 397514 (8.7%) |
| 60-65 | 182304 (7.9%) | 176855 (7.8%) | 359159 (7.9%) |
| 65-70 | 154461 (6.7%) | 146763 (6.5%) | 301224 (6.6%) |
| 70-75 | 141644 (6.1%) | 129545 (5.7%) | 271189 (5.9%) |
| 75-80 | 110509 (4.8%) | 93703 (4.2%) | 204212 (4.5%) |
| 80-85 | 76572 (3.3%) | 55910 (2.5%) | 132482 (2.9%) |
| 85-90 | 48220 (2.1%) | 29547 (1.3%) | 77767 (1.7%) |
| 90+ | 26797 (1.2%) | 11948 (0.5%) | 38745 (0.8%) |

### Simulation scenarios

We simulated one clinic visit, representing vaccine allocation during the study period. The treatment variable A_0_ ∈ {0,1}, was specified by logistic models with parameters γ estimated by fitting a logistic regression to aggregated EAVE-II vaccination counts (stratified by sex and age group, extracted August 2025). The outcome was modelled as a binary MSM (23)with ’ground truth’ parameter values reported in Table 2. The confounding mechanism was determined by the risk score function, with ’ground truth’ parameter values reported in Table 3.

**Table 2.** ‘Ground truth’ parameter values for the outcome model parameters β.

|  | Parameter | Thrombocytopenia | ITP | Venous | Arterial | Haemorrhagic |
| --- | --- | --- | --- | --- | --- | --- |
| Intercept | $\beta_0$ | -5.29 | -5.29 | -5.29 | -5.29 | -5.29 |
| $A_0 = 1$ | $\beta_1$ | 0.35 | 1.75 | 0.03 | 0.20 | 0.39 |

ITP = immune thrombocytopenic purpura; $\beta_0$ = intercept; $\beta_1$ = coefficient for first vaccine dose.

**Table 3.** ‘Ground truth’ parameter values for the risk score parameters θ.

|  | Parameter | 'Ground truth' |
| --- | --- | --- |
| Sex = M | $\theta_1$ | 2 |
| Age $\geq 15$ | $\theta_2$ | 1 |
| : | : | : |
| Age $\geq 90$ | $\theta_{16}$ | 1 |

$\theta$ = risk score parameters; M = male.

Five vaccine-safety-related rare adverse outcomes were considered: thrombocytopenia, immune thrombocytopenic purpura (ITP), venous thromboembolism, arterial thromboembolism, and haemorrhagic stroke (32). ‘Ground truth’ outcome parameters were informed by clinical events extracted from EAVE-II on 8 November 2025, stratified by event type, sex, age group, and vaccination status.

The strength of confounding was varied across five simulations with ρₖ ∈ {−0.1, −0.3, −0.5, −0.7, −0.9}. For each scenario, logistic regression models were fitted for each outcome using (a) no weighting and (b) Inverse Probability of Treatment Weighting (IPTW) (23).

## Results

Synthetic datasets of 100,000 individuals were generated under each of the five confounding scenarios and five outcome types. For each of those twenty-five scenario-outcome combinations, 1,000 synthetic datasets were generated. The baseline characteristics of the simulated population closely reflected the distribution of the Scottish population as captured in aggregated EAVE-II summaries (Table 1).

For weak confounding (ρₖ = −0.1), the true values of the parameters determining the causal mechanism were recovered when fitting both unweighted and weighted (IPTW) logistic regression models. As confounding strength increased (ρₖ decreasing toward −0.9), the unweighted models showed increasing bias and coverages far below the nominal 95%, indicating substantial confounding. By contrast, the IPTW-weighted models continued to recover the true causal parameters across a wide range of confounding strengths, though with some small residual bias as ρₖ approached its lower limit of −1. Full simulation results for all outcome types are provided in the supplementary material (Figure 2; Supplementary Tables S1–S5).

**Figure 2.**
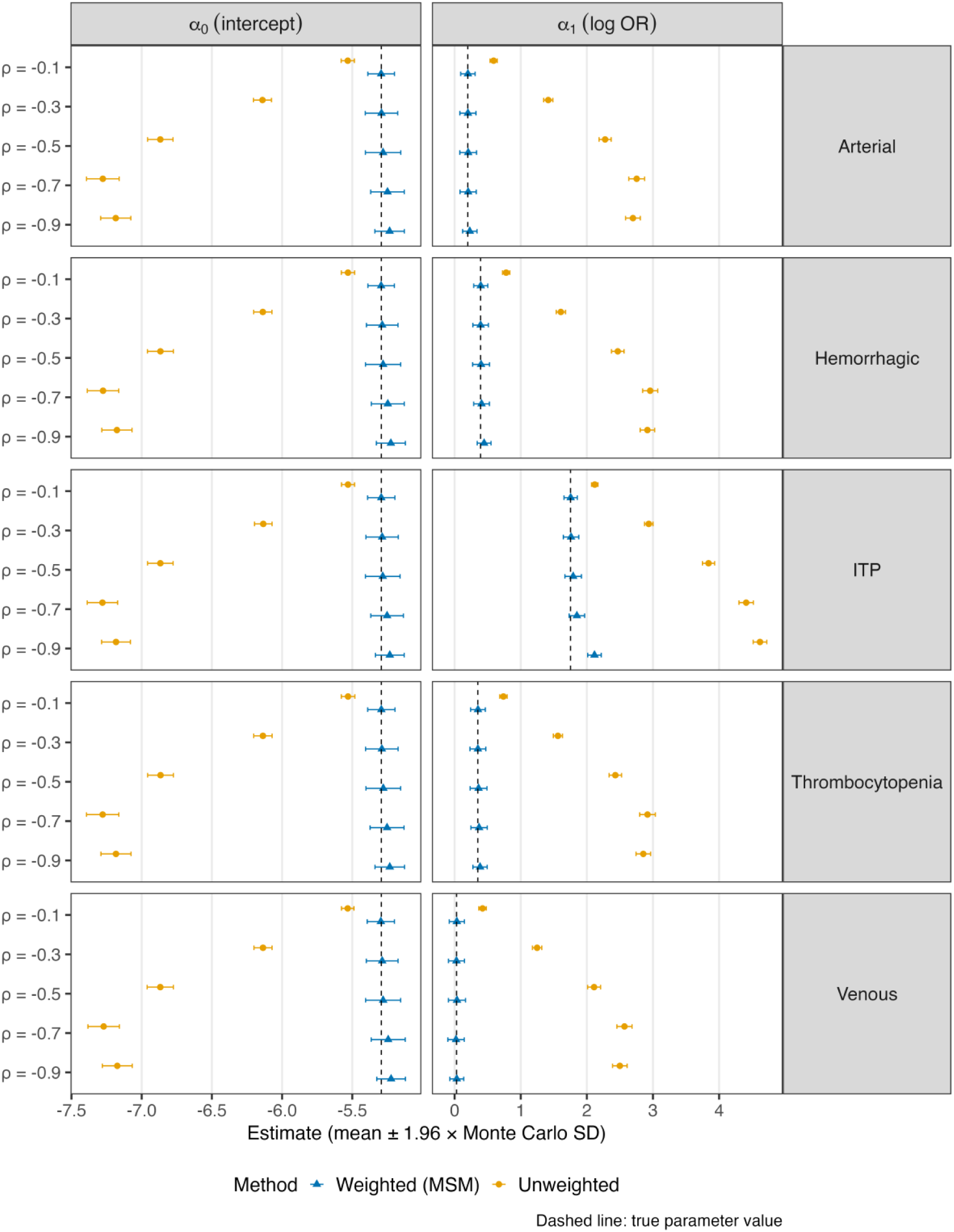
Unweighted and inverse probability weighted (IPW) estimates of both parameters for all five outcomes and all five scenarios. Points represent the mean estimate across simulation replicates; horizontal bars show ±1.96 Monte Carlo standard deviations. The dashed vertical line indicates the true parameter value.

The workflow enabled by synthetic data is illustrated in Figure 3. Under the standard EAVE-II workflow, analysts cannot proceed with analysis until data access has been formally approved, causing delays. The proposed alternative — using synthetic datasets parameterised from aggregated EAVE-II data — allows analysts to implement and test their full analysis pipeline in parallel with the access application process.

**Figure 3.**
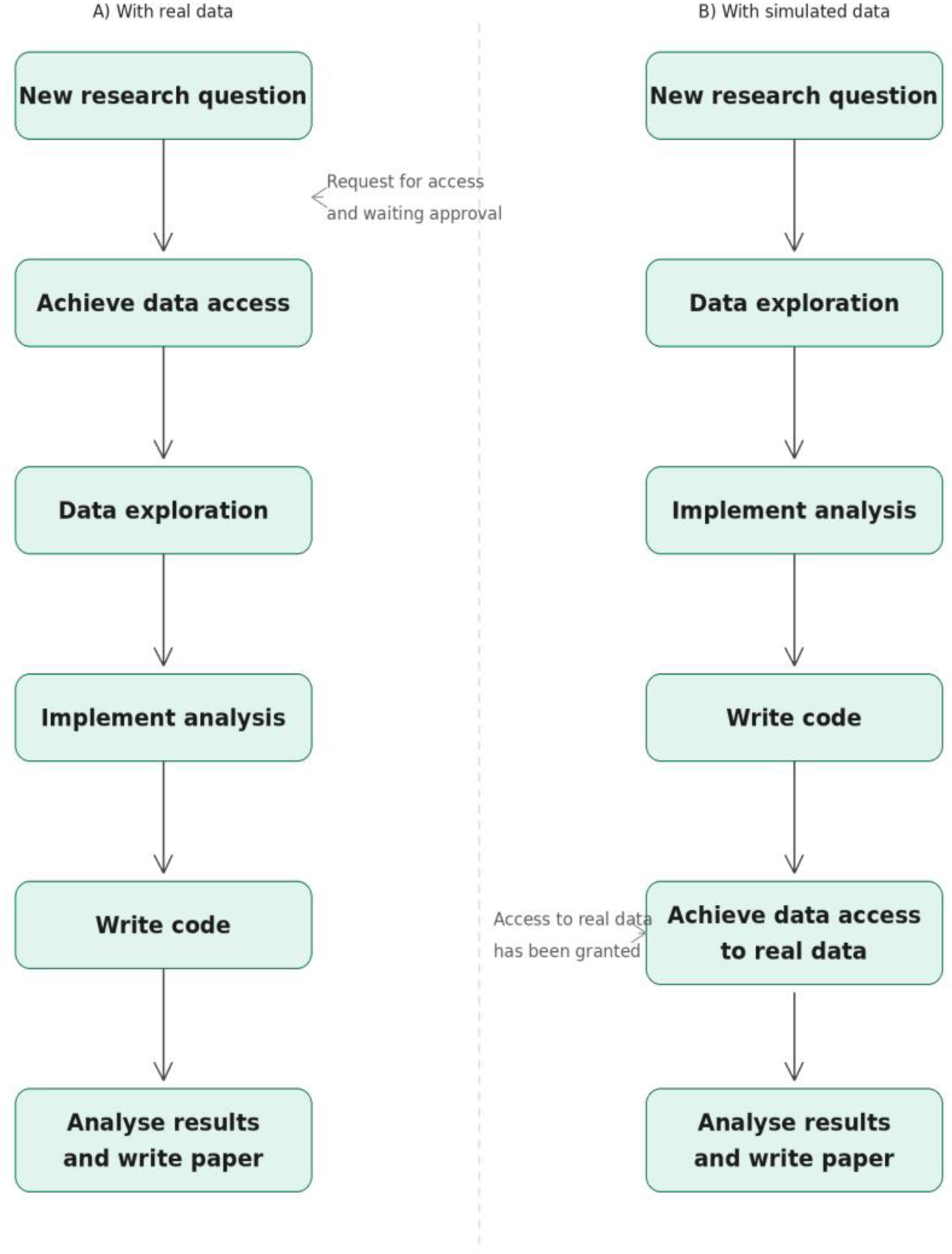
A) Typical workflow followed by an analyst in the EAVE-II team, in which progression is blocked until data access is approved. B) Suggested alternative workflow with simulated data, allowing analysis implementation and testing while awaiting access to real data.

## Discussion

We have generated synthetic health records that explicitly encode causal and confounding mechanisms, parameterised from aggregated EAVE-II data, using a statistically sound method grounded in the methodological literature. Our approach looks to address a key bottleneck in public health research whereby access to individual-level data for methodological development is constrained. By providing synthetic datasets in advance of or as a substitute to data access, analysts can develop, test, and assess causal inference pipelines whilst waiting for data access to be granted. Additionally, synthetic data of sufficient fidelity can enable training in carrying out causal inference for vaccine effectiveness research in a setting that closely mirrors real analyses.

We emphasise that these synthetic datasets are intended for pipeline development and training purposes only. They are not intended to answer clinical or public health questions: the data are parameterised from aggregated EAVE-II statistics and do not reproduce the individual-level structure of the underlying records, and therefore should not be used to draw clinical or policy conclusions.

The Seaman and Keogh framework(30), on which our implementation is based, offers important advantages over simpler joint distribution-based approaches: the causal mechanism is explicitly specified, and it is possible to directly verify that causal inference methods recover the true parameter values. However, the framework requires the practitioner to implement the simulation algorithm, as no readily available R package currently exists for this purpose.

The main limitation of our synthetic simulations is as follows. The simulation framework developed by Seaman and Keogh (30) is designed for settings in which treatment allocation is assessed at each visit and, within every covariate stratum, the probability of receiving treatment is strictly between 0 and 1. In principle, this supports simulations with multiple sequential follow-up visits. However, the EAVE-II analysis on which our simulation is based (32) reports outcomes at weekly intervals following a single vaccination dose. Because the minimum interval between vaccine doses is far longer than one week, a patient who has already received their first dose cannot receive another within the next weekly visit. Consequently, we were only able to simulate a scenario comprising a single baseline visit at which treatment is allocated and one subsequent visit at which the outcome is assessed. Future work will perform a more complex longitudinal simulation where multiple visits are considered.

## Conclusions

Data access applications remain a bottleneck for the development of novel causal inference methods and pipelines in public health research. We created synthetic datasets resembling data that could be egressed from the EAVE-II platform, using aggregated EAVE-II data extracted in 2025 to parameterise both treatment and outcome mechanisms. Building on the method of Seaman and Keogh (30), we were able to specify ’ground truth’ mechanisms for treatment allocation, causal effect of treatment on outcomes, and confounding effect of demographic variables. The synthetic datasets will allow the EAVE-II team and other practitioners to develop the pipelines needed to use causal inference methods while waiting for data access to be granted, prototype and evaluate new analytical approaches without requiring sensitive data and provide a training resource for analysts.

## Statements

### Ethics statement

Not applicable as only using publicly available and simulated data. Ethical approval for the original EAVE II study was granted by the National Research Ethics Service Committee (Southeast Scotland 02; reference number 12/SS/0201), and the required data linkage was approved by the Public Benefit and Privacy Panel for Health and Social Care (reference number 1920–0279).

### Data availability

Code to reproduce all results in this paper is available at https://github.com/victorvelasco/eave_ii_simulated_data. No individual-level patient data were used or generated; the aggregated EAVE-II summaries used to parameterise the simulation are described in the Methods section.

### Authorship contributions

Conceptualization: VVP, BS, CM, CS; Methodology: VVP; Software: VVP; Formal analysis: VVP; Investigation: VVP; Data curation: VVP; Writing – original draft: VVP; Writing – review & editing: BS, CM, CS, LD, LR Supervision: BS, CM; Project administration: BS, CM; Funding acquisition: BS, CM, CS.

## Acknowledgements

VVP would like to thank Dr. Shaun Seaman (MRC Biostatistics Unit, University of Cambridge) for kindly having shared the R code accompanying (30) prior to its publication, and Dr. Steven Kerr (Usher Institute, University of Edinburgh) for extracting and providing the aggregate data used to simulate the synthetic data.

## Appendix A: Data simulation algorithm

The following pseudocode reproduces the data simulation algorithm of Seaman and Keogh (30). We iterate the following steps a number of times equal to the desired sample size:

1. Sample Z from p(Z) and set k = 0.
2. Sample Aₖ from p(Aₖ|Z, Ā_{k−1}, Yₖ = 1).
3. Calculate h(Z, Āₖ).
4. Sample U_{H,k,Āₖ} from F_H(h(Z, Āₖ)).
5. Calculate Z_{H,k,Āₖ}.
6. Sample Z_{Y,k,Āₖ} from N_1_(ρₖ · Z_{H,k,Āₖ}, 1 − ρₖ²) and calculate U_{Y,k,Āₖ} = Φ(Z_{Y,k,Āₖ}).
7. If U_{Y,k,Āₖ} ≤ P*(Yₖ₊_1_ = 0|Āₖ), set Yₖ₊_1_ = 0, else set Yₖ₊_1_ = 1.
8. If Yₖ₊_1_ = 1 and k < K, set k = k + 1 and go to step 2.

F_H denotes the CDF of random variable H; N_1_ denotes the standard normal distribution; Φ is its CDF; U denotes the continuous uniform distribution.

## Appendix B: Supplementary tables

**Table S1.**
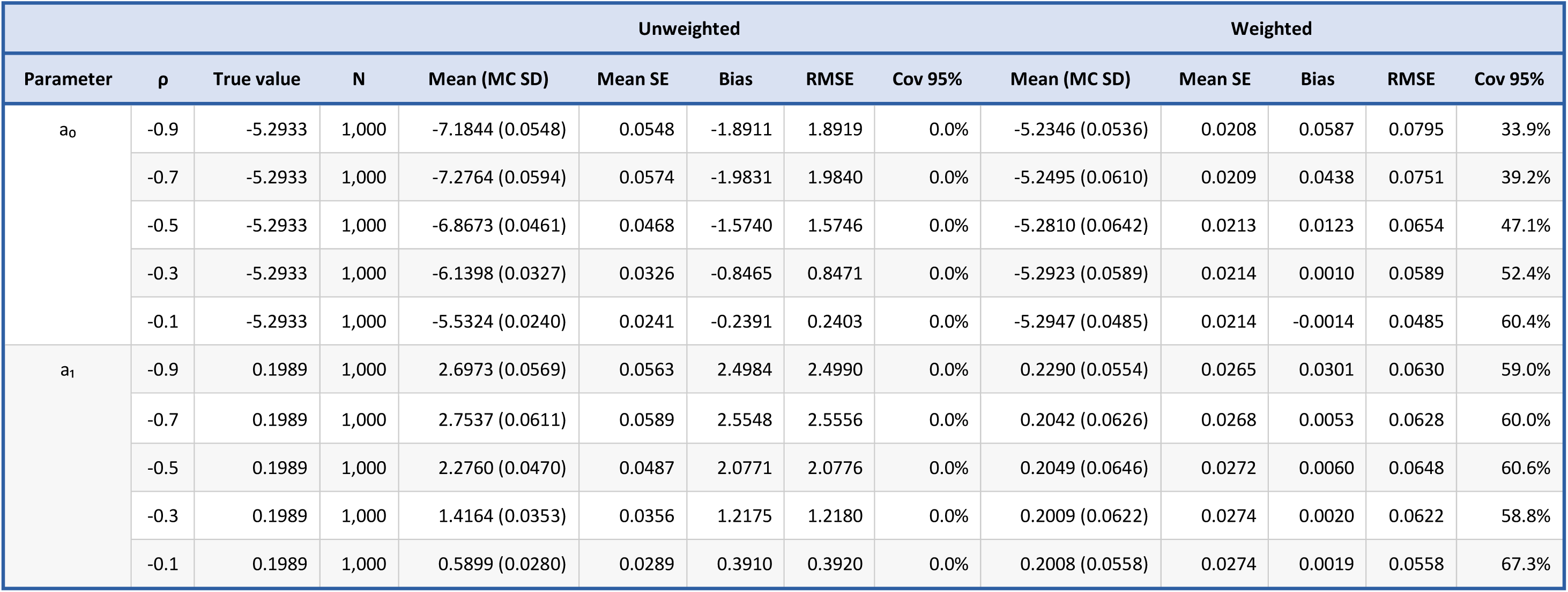
Simulation results (unweighted and weighted) for the Arterial outcome. Based on 1000 simulations per cell.

**Table S2.** Simulation results (unweighted and weighted) for the Haemorrhagic outcome. Based on 1000 simulations per cell.

| Parameter | $\rho$ | True value | N | Unweighted | | | | | Weighted | | | | |
| --- | --- | --- | --- | --- | --- | --- | --- | --- | --- | --- | --- | --- | --- |
|  |  |  |  | Mean (MC SD) | Mean SE | Bias | RMSE | Cov 95% | Mean (MC SD) | Mean SE | Bias | RMSE | Cov 95% |
| $a_0$ | -0.9 | -5.2933 | 1,000 | -7.1759 (0.0549) | 0.0546 | -1.8826 | 1.8834 | 0.0% | -5.2257 (0.0527) | 0.0207 | 0.0676 | 0.0857 | 29.3% |
|  | -0.7 | -5.2933 | 1,000 | -7.2749 (0.0571) | 0.0574 | -1.9816 | 1.9824 | 0.0% | -5.2476 (0.0599) | 0.0209 | 0.0457 | 0.0754 | 41.1% |
|  | -0.5 | -5.2933 | 1,000 | -6.8664 (0.0471) | 0.0468 | -1.5731 | 1.5738 | 0.0% | -5.2810 (0.0639) | 0.0213 | 0.0123 | 0.0650 | 48.1% |
|  | -0.3 | -5.2933 | 1,000 | -6.1374 (0.0331) | 0.0325 | -0.8441 | 0.8447 | 0.0% | -5.2872 (0.0574) | 0.0213 | 0.0061 | 0.0577 | 53.8% |
|  | -0.1 | -5.2933 | 1,000 | -5.5309 (0.0241) | 0.0241 | -0.2376 | 0.2388 | 0.0% | -5.2949 (0.0483) | 0.0214 | -0.0016 | 0.0483 | 62.3% |
| $a_1$ | -0.9 | 0.3920 | 1,000 | 2.9163 (0.0559) | 0.0558 | 2.5243 | 2.5249 | 0.0% | 0.4457 (0.0536) | 0.0254 | 0.0537 | 0.0758 | 47.0% |
|  | -0.7 | 0.3920 | 1,000 | 2.9580 (0.0582) | 0.0586 | 2.5660 | 2.5666 | 0.0% | 0.4071 (0.0608) | 0.0258 | 0.0151 | 0.0627 | 59.3% |
|  | -0.5 | 0.3920 | 1,000 | 2.4676 (0.0486) | 0.0484 | 2.0756 | 2.0762 | 0.0% | 0.3999 (0.0651) | 0.0263 | 0.0079 | 0.0656 | 57.7% |
|  | -0.3 | 0.3920 | 1,000 | 1.6067 (0.0357) | 0.0350 | 1.2147 | 1.2152 | 0.0% | 0.3930 (0.0599) | 0.0264 | 0.0010 | 0.0599 | 61.3% |
|  | -0.1 | 0.3920 | 1,000 | 0.7797 (0.0281) | 0.0281 | 0.3877 | 0.3887 | 0.0% | 0.3951 (0.0546) | 0.0265 | 0.0031 | 0.0547 | 66.4% |

**Table S3.** Simulation results (unweighted and weighted) for the ITP outcome. Based on 1000 simulations per cell.

| Parameter | $\rho$ | True value | N | Unweighted | | | | | Weighted | | | | |
| --- | --- | --- | --- | --- | --- | --- | --- | --- | --- | --- | --- | --- | --- |
|  |  |  |  | Mean (MC SD) | Mean SE | Bias | RMSE | Cov 95% | Mean (MC SD) | Mean SE | Bias | RMSE | Cov 95% |
| $a_0$ | -0.9 | -5.2933 | 1,000 | -7.1824 (0.0523) | 0.0548 | -1.8891 | 1.8898 | 0.0% | -5.2331 (0.0523) | 0.0208 | 0.0602 | 0.0797 | 34.9% |
|  | -0.7 | -5.2933 | 1,000 | -7.2791 (0.0552) | 0.0575 | -1.9858 | 1.9866 | 0.0% | -5.2524 (0.0592) | 0.0210 | 0.0409 | 0.0720 | 42.5% |
|  | -0.5 | -5.2933 | 1,000 | -6.8671 (0.0464) | 0.0468 | -1.5738 | 1.5745 | 0.0% | -5.2833 (0.0629) | 0.0213 | 0.0100 | 0.0637 | 46.6% |
|  | -0.3 | -5.2933 | 1,000 | -6.1341 (0.0319) | 0.0325 | -0.8408 | 0.8414 | 0.0% | -5.2877 (0.0588) | 0.0213 | 0.0056 | 0.0590 | 53.2% |
|  | -0.1 | -5.2933 | 1,000 | -5.5306 (0.0237) | 0.0241 | -0.2373 | 0.2384 | 0.0% | -5.2939 (0.0496) | 0.0214 | -0.0006 | 0.0496 | 61.5% |
| $a_1$ | -0.9 | 1.7527 | 1,000 | 4.6187 (0.0526) | 0.0550 | 2.8660 | 2.8664 | 0.0% | 2.1151 (0.0523) | 0.0218 | 0.3624 | 0.3662 | 0.0% |
|  | -0.7 | 1.7527 | 1,000 | 4.4099 (0.0556) | 0.0578 | 2.6572 | 2.6578 | 0.0% | 1.8490 (0.0595) | 0.0223 | 0.0963 | 0.1132 | 18.6% |
|  | -0.5 | 1.7527 | 1,000 | 3.8406 (0.0470) | 0.0472 | 2.0879 | 2.0884 | 0.0% | 1.7929 (0.0636) | 0.0227 | 0.0402 | 0.0752 | 44.3% |
|  | -0.3 | 1.7527 | 1,000 | 2.9362 (0.0328) | 0.0332 | 1.1835 | 1.1840 | 0.0% | 1.7608 (0.0594) | 0.0228 | 0.0081 | 0.0600 | 55.0% |
|  | -0.1 | 1.7527 | 1,000 | 2.1196 (0.0246) | 0.0252 | 0.3669 | 0.3677 | 0.0% | 1.7550 (0.0512) | 0.0229 | 0.0023 | 0.0513 | 64.4% |

**Table S4.** Simulation results (unweighted and weighted) for the Thrombocytopenia outcome. Based on 1000 simulations per cell.

| Parameter | $\rho$ | True value | N | Unweighted | | | | | Weighted | | | | |
| --- | --- | --- | --- | --- | --- | --- | --- | --- | --- | --- | --- | --- | --- |
|  |  |  |  | Mean (MC SD) | Mean SE | Bias | RMSE | Cov 95% | Mean (MC SD) | Mean SE | Bias | RMSE | Cov 95% |
| $a_0$ | -0.9 | -5.2933 | 1,000 | -7.1828 (0.0547) | 0.0548 | -1.8895 | 1.8903 | 0.0% | -5.2334 (0.0538) | 0.0208 | 0.0599 | 0.0805 | 33.2% |
|  | -0.7 | -5.2933 | 1,000 | -7.2776 (0.0589) | 0.0575 | -1.9843 | 1.9852 | 0.0% | -5.2526 (0.0616) | 0.0210 | 0.0407 | 0.0739 | 41.1% |
|  | -0.5 | -5.2933 | 1,000 | -6.8656 (0.0468) | 0.0468 | -1.5723 | 1.5730 | 0.0% | -5.2794 (0.0629) | 0.0213 | 0.0139 | 0.0644 | 49.3% |
|  | -0.3 | -5.2933 | 1,000 | -6.1366 (0.0333) | 0.0325 | -0.8433 | 0.8440 | 0.0% | -5.2897 (0.0591) | 0.0214 | 0.0036 | 0.0592 | 52.3% |
|  | -0.1 | -5.2933 | 1,000 | -5.5303 (0.0245) | 0.0241 | -0.2370 | 0.2383 | 0.0% | -5.2933 (0.0497) | 0.0214 | 0.0000 | 0.0496 | 60.8% |
| $a_1$ | -0.9 | 0.3507 | 1,000 | 2.8537 (0.0561) | 0.0560 | 2.5030 | 2.5037 | 0.0% | 0.3845 (0.0552) | 0.0257 | 0.0338 | 0.0647 | 54.9% |
|  | -0.7 | 0.3507 | 1,000 | 2.9179 (0.0605) | 0.0587 | 2.5672 | 2.5679 | 0.0% | 0.3697 (0.0632) | 0.0261 | 0.0190 | 0.0659 | 55.8% |
|  | -0.5 | 0.3507 | 1,000 | 2.4307 (0.0481) | 0.0484 | 2.0800 | 2.0806 | 0.0% | 0.3618 (0.0648) | 0.0264 | 0.0111 | 0.0657 | 58.1% |
|  | -0.3 | 0.3507 | 1,000 | 1.5625 (0.0353) | 0.0351 | 1.2118 | 1.2123 | 0.0% | 0.3515 (0.0610) | 0.0266 | 0.0008 | 0.0610 | 59.3% |
|  | -0.1 | 0.3507 | 1,000 | 0.7382 (0.0290) | 0.0282 | 0.3875 | 0.3886 | 0.0% | 0.3527 (0.0568) | 0.0267 | 0.0020 | 0.0568 | 64.0% |

**Table S5.**
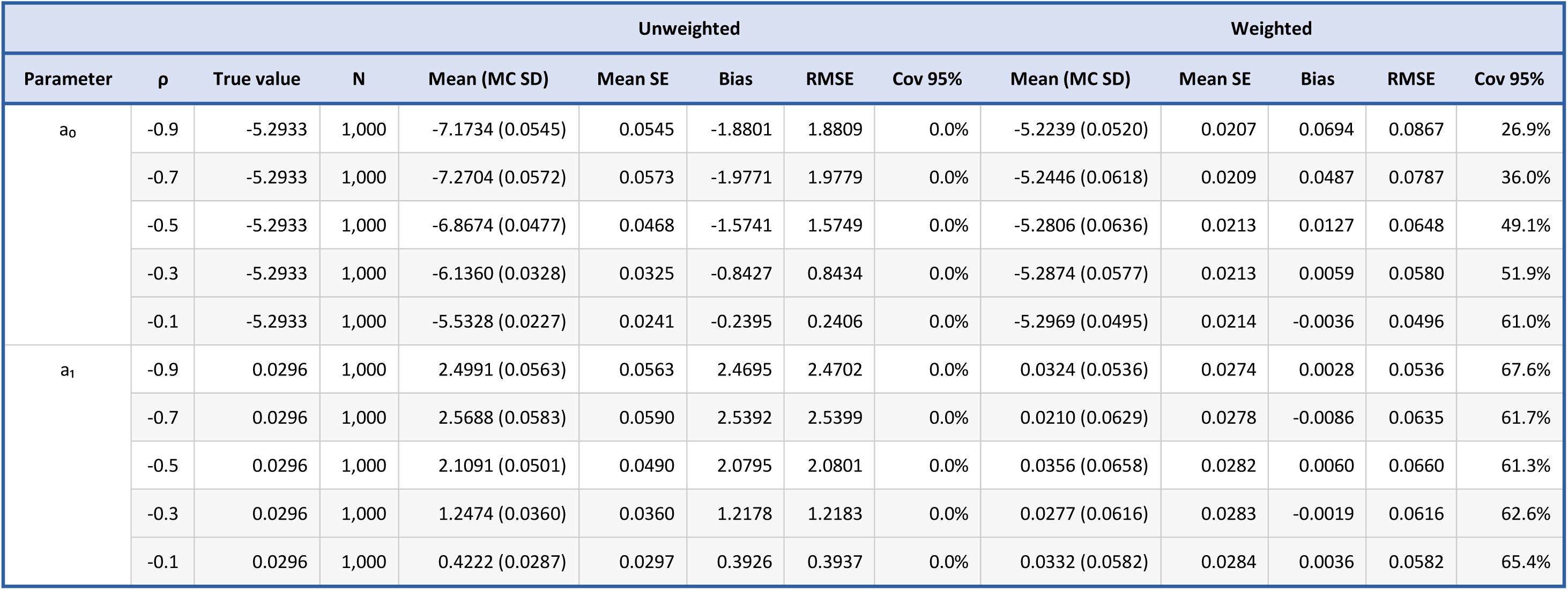
Simulation results (unweighted and weighted) for the Venous outcome. Based on 1000 simulations per cell.

